# Analysis on the Clinical Characteristics of 36 Cases of Novel Coronavirus Pneumonia in Kunming

**DOI:** 10.1101/2020.02.28.20029173

**Authors:** Haiyan Fu, Hongjuan Li, Xiaoqing Tang, Xiang Li, Jie Shen, Yujun Zhou, Bing Xu, Yu Luo

## Abstract

**Objective:** To analyze the clinical characteristics of patients with novel coronavirus pneumonia in Kunming City, and to study the correlation between nutritional status and immune function.

**Methods:** Clinical data of 36 patients with novel coronavirus pneumonia in isolation area of Kunming Third People’s Hospital from January 31 to February 15, 2020 were collected, and the basic situation, clinical characteristics, laboratory examination and CT imaging characteristics were analyzed. Serum albumin (ALB), prealbumin (PAB), hypersensitive c-reactive protein (hs-crp), CD3T cells, CD4T cells, CD8T cells and normal control group were analyzed. A simple linear regression analysis of the relationship between proalbumin and T cell subpopulation counts in the blood of patients.

**Results:** (1) The patients with new coronavirus pneumonia in Kunming were mainly of common type. (2) 50% of the patients’ first symptoms were fever and cough; (3) The total number of white blood cells in peripheral blood was normal or decreased in 23 cases (79%), and the lymphocyte count decreased in 5 cases (13.89%), without anemia. Hypersensitive c-reactive protein increased in 19 (52.78%) cases, and procalcitonin increased in 1 case. Albumin decreased in 5 cases (13.89%), proalbumin decreased in 15 cases (41.67%), alanine transaminase increased slightly in 4 cases (11.11%), alanine transaminase increased slightly in 4 cases (11.11%), total bilirubin increased slightly in 11 cases (30.56%), and renal function and blood coagulation were normal. Absolute value of CD3^+^T cells is with a decrease in 21 cases (58.3%), CD4^+^T in 28 cases (77.8%), CD8^+^T in 17 cases (47.2%), and CD4^+^/ CD8^+^ inverse in 6 cases (16.7%). (4) The prealbumin, CD3 T cells, CD4 T cells and CD8 T cells in the new coronavirus pneumonia group were significantly lower than those in the normal control group, and the hypersensitive c-reactive protein was higher than that in the normal control group. (5) The levels of PAB in the serum of the patients were linearly correlated with hs-crp, CD3 T cells, CD4 T cells and CD8 T cells, and the correlation coefficients were −0.474, 0.558, 0.467 and 0.613, respectively, showing statistical differences.

**Conclusion:** The clinical characteristics of the novel coronavirus pneumonia in Kunming are different from those in Wuhan. The changes of serum proalbumin and T cell subsets are relatively obvious. Changes in serum proalbumin may contribute to the early warning of novel coronavirus pneumonia. The nutritional status of patients with common and mild pneumonia should be considered.

## INTRODUCTION

In December 2019, a new outbreak of coronavirus pneumonia began in Wuhan, Hubei Province, and it quickly spread to the whole country, even many overseas countries. According to the outbreak dynamics released by China, most of the cases have come from Hubei, but outbreaks have been detected throughout the country. As a place famous for tourism, Yunnan has a large number of imported cases, and the incidence level is in the middle of the whole country. By the time of 24 o’clock on 15 February, a total of 169 cases were confirmed in Yunnan, including 49 in Kunming City (8 cured and discharged). Recently, there have been a large number of articles on novel coronavirus pneumonia, involving virus sequencing, clinical analysis and epidemiological studies. Among them, the disease model of epidemic studies is used to predict more. Those articles are mainly based on the data from Hubei, and there are few clinical data from other imported areas. The Third People’s Hospital of Kunming, as a designated hospital for the treatment of the new coronavirus pneumonia in Kunming, admitted about 3/4 of the patients diagnosed with the novel coronavirus pneumonia in Kunming. Through clinical analysis, it is found that the clinical characteristics of the pneumonia in Kunming are not exactly the same as the data reported in Wuhan. The hospitalized patients in Kunming are mainly of normal type and light type, and a small number of patients with severe or critical type recovered relatively well. By 24:00 on February 17, 2020, there were no death cases. In our clinical work, we found significant changes in patients’ nutritional status, which may be related to inflammation and immunity. This paper will analyze the clinical characteristics of the new type of coronavirus pneumonia admitted to the Third People’s Hospital of Kunming City, and analyze the relationship between serum proalbumin and T cells and its clinical significance.

### 1. Objects and Methods

#### 1.1 Research Objects

This study was approved by the ethics committee of The Third People’s Hospital of Kunming, and all data were used for clinical studies only, with the written consent of patients and their guardians. According to the diagnosis and treatment scheme issued by the National Health Commission of China, “the new coronavirus infection pneumonia diagnosis and treatment plan (trial edition5), the diagnostic criteria for new coronavirus pneumonia confirmed cases outside of Hubei province^[1]^: (1) The history of epidemiology: 1) Within 14 days before the onset, there was a history of traveling or living in the community of Wuhan city, surrounding areas, or other pathological reported area ^[1]^ 2) Within 14 days before the onset, there was contact history with cases of the new coronavirus infection (positive nucleic acid detection); 3) Within 14 days before the onset, there was contact with the patient with fever or respiratory symptoms from Wuhan, surrounding areas or from communities where cases of the disease have been reported; 4) Aggregation of the disease. (2) Clinical manifestations: 1) fever and/or respiratory symptoms; 2) With the above imaging characteristics of pneumonia; 3) The total number of early-onset white blood cells is normal or decreased, or lymphocyte count decreased. Any case is with 1 condition in the epidemiological history and conforms to any 2 in the clinical manifestations, or if there is no clear history of epidemiology, and it conforms to 3 clinical manifestations, the case can be diagnosed as suspected. Cases were included as suspected cases with one of the following etiological evidence:(1) Real-time fluorescence rt-pcr of respiratory tract specimens or hematological specimens was positive for new coronavirus nucleic acid; (2) Viral genetic sequencing of respiratory or hematological specimens, is highly homologous with known novel coronaviruses. Futhermore, patients were included in the inclusion criteria, when the following 2 criteria were met: (1) Suspected cases of new coronavirus pneumonia; (2) Real-time fluorescence rt-pcr was performed on respiratory specimens to detect positive nucleic acid of novel coronavirus. Clinical data were collected from 36 patients with novel coronavirus pneumonia who met the criteria above in the isolation area of Kunming Third People’s hospital from January 26 to February 15, 2020. The normal control group came from healthy people in the physical examination center.

#### 1.2. Clinical Classification of Disease Severity

On admission, the diagnosed patients were clinically classified according to the “pneumonia diagnosis and treatment plan for new coronavirus infection (trial version 5)”, as follows: (1) Mild type: with mild clinical symptoms, and without imaging finding of pneumonia (2) Common type: with fever, respiratory tract and other symptoms, and with imaging finding of pneumonia; (3) Severe type: accord with any of the following: 1) Respiratory distress, respiratory rate (RR) ≥30 times/min; 2) In resting state, oxygen saturation ≤93%; 3) Partial arterial oxygen pressure (PaO)/oxygen absorption concentration (FiO) ≤300mmHg (1mmHg= 0.133kpa);^22^ (4) Critical type: meet one of the following conditions: 1) Respiratory failure occurs, and mechanical ventilation is required; 2) Shock;3) Complicated with other organ failure and requiring intensive care of ICU unit.

#### 1.3. Statistical processing

SPSS 20.0 software was used for data analysis. Case or rate were used for data counting, and χ2 test was used for inter-group comparison; The measurement data of normal distribution were expressed as x±s, and the measurement data of non-normal distribution were expressed as median (quartile spacing). The measurement data of normal distribution was verified with t-test or analysis of variance, and the rank sum test was used for inter-group comparison of non-normal distribution. The relationship between PAB and T cell subsets and hs-crp was analyzed by regression, and the scatter plot was made by simple linear regression and the regression equation was calculated. P < 0.05 was considered statistically significant.

## 2. Results

### 2.1 clinical features: general data

36 patients were diagnosed at the age of 3-79 years, with a median age of 45 years, including 4 children (11.11%). There were 16 males (44.44%) and 20 females (55.56%). There were 21 cases with long-term exposure, 12 cases with short-term exposure, and 3 cases with unknown exposure time. There were 10 previous smoking cases (27.78%). There were 10 patients with basic diseases (27.78%), including 7 patients with hypertension (19.44%), 4 patients with diabetes (11.11%), 1 patient with hypothyroidism (2.78%), and 1 patient with hyperlipidemia (2.78%). There were 4 cases of light type, 30 cases of normal type and 2 cases of critical type. There were 18(50%) cases in Hubei and 18(50%) cases in Yunnan.

Signs and symptoms(table 1): of the 36 patients confirmed,fever in 18 cases (50%), cough in 18 cases (50%), expectoration in 11 cases (30.56%), shortness of breath in 1 case (2.78%), runny nose in 8 cases (22.22%), blood sputum in 1 case (2.78%), diarrhea in 3 patients (8.33%), pharynx pain in 6 cases (16.67%), dry throat in 2 cases (5.56%), chest pain in 1 case (2.78%), chest tightness in 3 patients (8.33%), muscle soreness in 6 cases (16.67%), headache in2 cases (5.56%), and lack of power in 3 patients (8.33%).

**Table 1:** Symptoms of 36 confirmed patients

| symptom | Number of cases | Proportion (%) |
| --- | --- | --- |
| heat | 18 | 50 |
| cough | 18 | 50 |
| Expectorant | 11 | 30.56 |
| Shortness of breath | 1 | 2.78 |
| Runny | 8 | 22.22 |
| Sputum | 1 | 2.78 |
| diarrhea | 3 | 8.33 |
| Sore throat | 6 | 16.67 |
| Dry throat | 2 | 5.56 |
| Chest pain | 1 | 2.78 |
| Chest tightness | 3 | 8.33 |
| Muscle ache | 6 | 16.67 |
| headache | 2 | 5.56 |
| Fatigue | 3 | 8.33 |

**Table 2:**
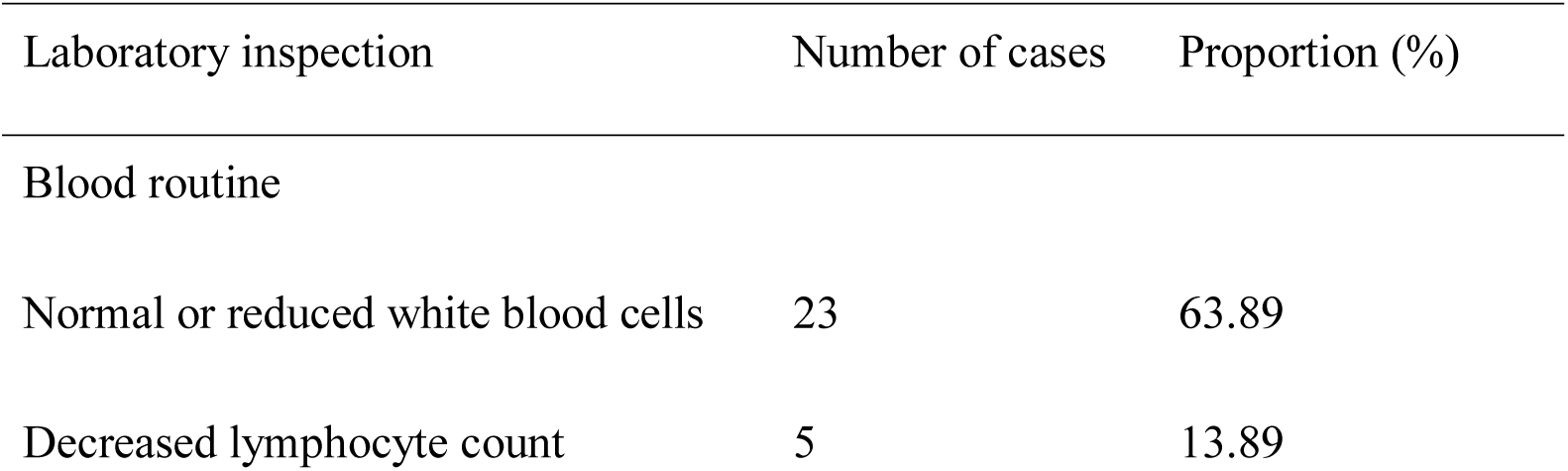
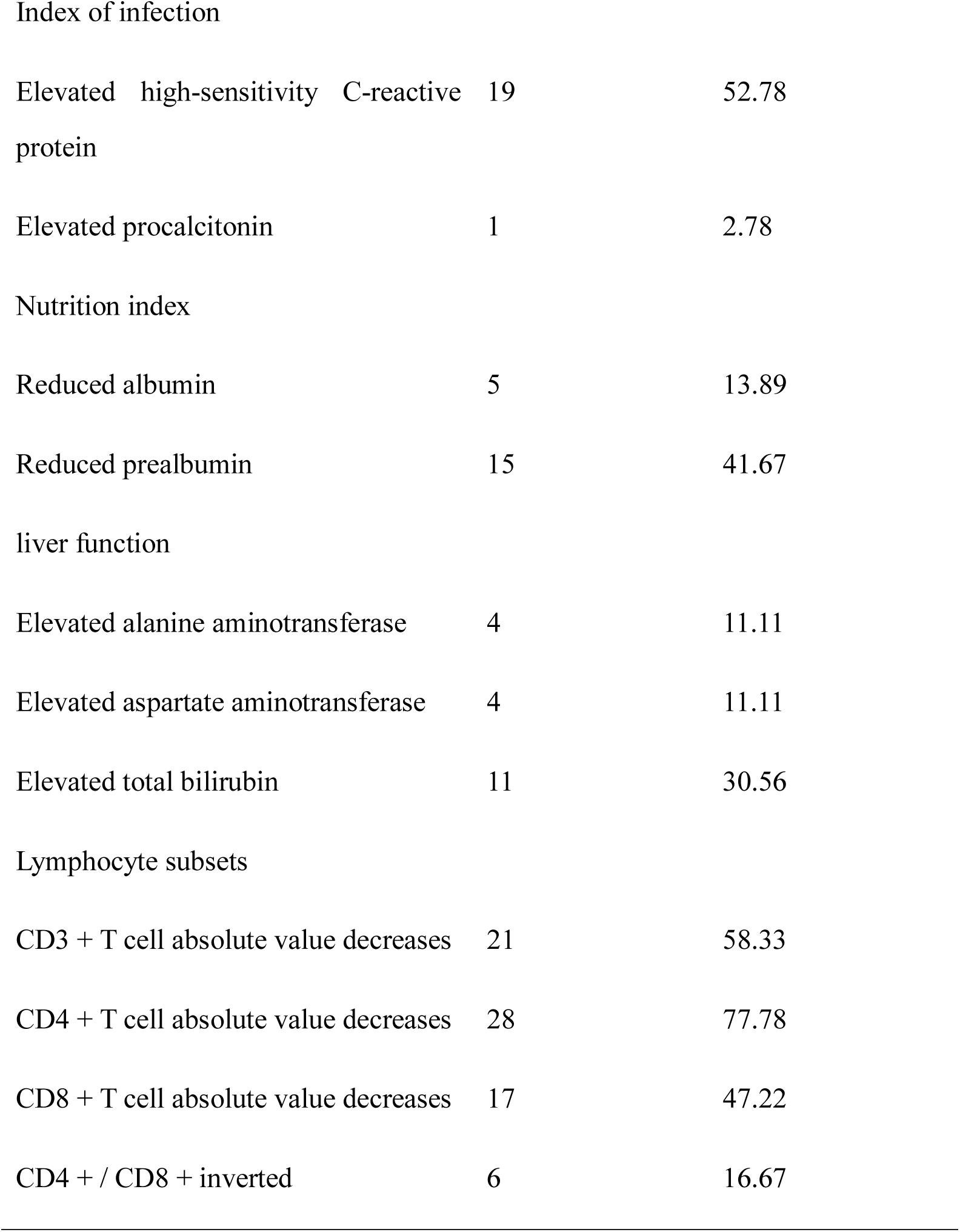
Laboratory examination of 36 confirmed patients

| Laboratory inspection | Number of cases | Proportion (%) |
| --- | --- | --- |
| Blood routine |  |  |
| Normal or reduced white blood cells | 23 | 63.89 |
| Decreased lymphocyte count | 5 | 13.89 |
| Index of infection |  |  |
| Elevated high-sensitivity C-reactive protein | 19 | 52.78 |
| Elevated procalcitonin | 1 | 2.78 |
| Nutrition index |  |  |
| Reduced albumin | 5 | 13.89 |
| Reduced prealbumin | 15 | 41.67 |
| liver function |  |  |
| Elevated alanine aminotransferase | 4 | 11.11 |
| Elevated aspartate aminotransferase | 4 | 11.11 |
| Elevated total bilirubin | 11 | 30.56 |
| Lymphocyte subsets |  |  |
| CD3 + T cell absolute value decreases | 21 | 58.33 |
| CD4 + T cell absolute value decreases | 28 | 77.78 |
| CD8 + T cell absolute value decreases | 17 | 47.22 |
| CD4 + / CD8 + inverted | 6 | 16.67 |

### 2.2 Laboratory examination (table 2)

Among the 36 confirmed patients, 23 (79%) had normal or decreased total white blood cells in peripheral blood, 5 (13.89%) had decreased lymphocyte count, and none had anemia. Hypersensitive c-reactive protein increased in 19 (52.78%) cases, and procalcitonin increased in 1 case. Albumin decreased in 5 cases (13.89%), prealbumin decreased in 15 cases (41.67%), alanine aminotransferase increased slightly in 4 cases and glutamic-oxalacetic transaminease increased slightly in 4 cases. Absolute value of CD3^+^T cells decreased in 21 cases (58.3%), CD4^+^T in 28 cases (77.8%), CD8^+^T in 17 cases (47.2%), and CD4^+^ /CD8^+^T inverse in 6 cases (16.7%).

### 2.3 CT imaging examination analysis(table3 and figure1)

Thirty-six patients were admitted for CT examination, of which 14 (38.89%) were negative and 22 (61.11%) were positive, involving only 2 (5.56%) of the left lung, 4 (11.11%) of the right lung, and 16 (44.44%) of the double lung. Pulmonary ground glass shadow was found in 22 (61.11%) cases (FIG. 1A), pavement stone sign in 14 (38.89%) cases (FIG. 1B), strip shadow in 10 (27.78%) cases (FIG. 1C), consolidation shadow in 7 (19.44%) cases (FIG. 1D), aerated bronchi sign in 4 (11.11%) cases (FIG. 1D), and vascular thickening in 7 (19.44%) cases (FIG. 1E). Typical performance is shown in figure 1:

**FIG. 1.**
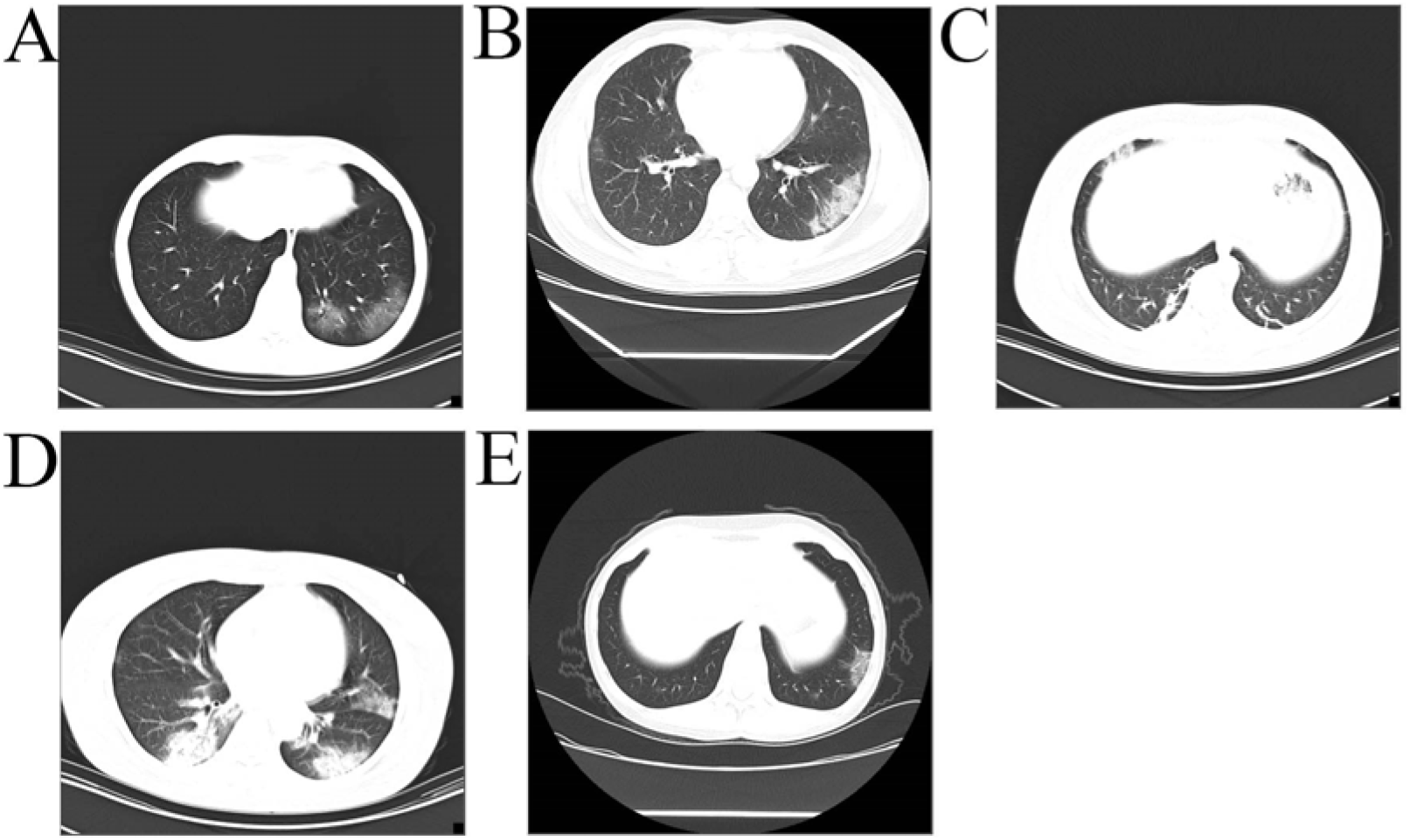
A. Large ground glass like shadows in the lower lobe and posterior basal segment of the left lung; B. the grid-shaped interlobular septa in the basal segment of the lower lobe of the left lung thickened, presenting a “paving stone” -like change; C. Multiple striated shadows in the posterior basal segment of the lower lobe of both lungs; D. Large consolidation shadow in the posterior basal segment of the lower lobe of the right lung. Ground glass and consolidation in the lower lobe and posterior basal segment of the left lung; E. ground glass shadow in the basal segment of the lower lobe of the left lung, and thickened blood vessels in the lesion

### 2.4 Treatment

With National Health Commission of China updating the new coronavirus infection pneumonia diagnosis and treatment scheme (trial version 5) for reference, all hospitalized patients were given Lopinavir and Ritonavir tablets for antiviral treatment, 7 cases of patients were with plus ribavirin therapy, 36 patients were all given oxygen therapy and interferon treatment, 32 cases patients were with antibiotic therapy, 28 patients were given Chinese medicine medicinal broth treatment (Small Bupleurum decoction, Wendan conditioning decoction, Buzhong Yiqi tonic decoction), 11 patients were with treatment to enhance immunity, 14 patients were with the using of immunoglobulin, 14 patients were with hormone therapy, of whom the treatment for patients with hypertension were altered to calcium antagonist, and 2 patients were treated with non-invasive ventilator assisted breathing.

### 2.5 Outcomes

34 patients were admitted to the general isolation wards, and 2 patients were admitted to the ICU isolation wards, During the treatment period, 1 patient’s condition worsened and was transferred to ICU for treatment. By 24:00 on February 17, 2020, 6 patients were discharged according to the discharge criteria of “pneumonia diagnosis and treatment program for novel coronavirus infection (trial version 5)”; Two patients were admitted to the ICU One of the two patients admitted to ICU got better on the 17^th^ day after admission, getting out of the critical condition patient was in critical condition on the 17th day after admission, and the other one patient, with stable condition, needs to be treated continuously,. The patients transferred to ICU are in stable condition. The rest are still in hospital and in stable condition.

### 2.6 Serum

albumin (ALB), prealbumin (PAB), hypersensitive c-reactive protein (hs-crp), CD3^+^T cells, CD4^+^T cells, CD8^+^T cells and normal control group were compared, as shown in table 4:

**Table 3:** CT imaging results of 36 confirmed patients

| Result | Number of cases | Proportion (%) |
| --- | --- | --- |
| Positive | 22 | 61.11 |
| One lung | 6 | 16.67 |
| Left lung | 2 | 5.56 |
| Right lung | 4 | 11.11 |
| Two lungs | 16 | 44.44 |
| negative | 14 | 38.89 |
| CT manifestation |  |  |
| Ground glass | 22 | 61.11 |
| Paving stone sign | 14 | 38.89 |
| Strip shadow | 10 | 27.78 |
| Real Shadow | 7 | 19.44 |
| Inflatable bronchi sign | 4 | 11.11 |
| Vascular thickening | 7 | 19.44 |

**Table 4:** results of ALB, PA, hs-crp and T cell subsets in serum of 36 patients. ALB: albumin; PAB: prealbumin; Hs-crp: hypersensitive c-reactive protein; * p < 0.05

|  | n | ALB<br>( g/L ) | PAB<br>( mg/L ) | Hs-CRP<br>( mg/L ) | CD3+<br>(A / ul) | CD4+<br>(A / ul) | CD8+<br>(A / ul) |
| --- | --- | --- | --- | --- | --- | --- | --- |
| Control | 25 | 40.57<br>±3.80 | 281.76<br>±61.46 | 2.68<br>±1.54 | 1400.84<br>±254.99 | 830.07<br>±172.95 | 462.32<br>±137.67 |
| COVID-19 | 36 | 39.98<br>±8.37 | 221.78<br>±69.66 | 5.84<br>±5.16 | 915.80<br>±401.81 | 528.36<br>±265.40 | 337.86<br>±152.20 |

The results show that the PAB, CD^+^3, CD^+^4 and CD^+^8 T cells in the pneumonia group of the novel coronavirus are significantly lower than those in the normal control group, and the hypersensitive c-reactive protein is higher than that in the normal control group, while there is no significant differences between the albumin and the normal control group.

Simple linear regression analysis of serum PAB, hs-crp, CD3^+^T cells, CD4 ^+^T cells and CD8^+^T cells in 36 patients with novel coronavirus pneumonia:

The level of PAB in the serum of the patients is linearly correlated with CD3^+^T cells, and the level of PAB in the serum of the patients is positively correlated with CD3^+^T cells, with a correlation coefficient R=0.558. After T test of PAB, T =3.92, p=0.000, it shows a statistical difference (figure 2B).

**Figure 2.**
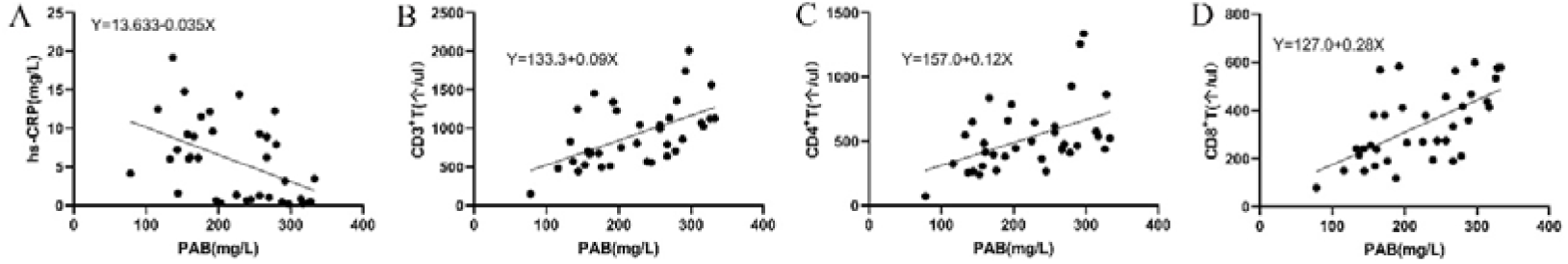
changes of serum PAB and a. hs-crp in 36 patients with novel coronavirus pneumonia; B. CD3^+^T cells; C. CD4^+^T cells; D. linear relationship of CD8^+^T cells The serum PAB level of the patients is linearly correlated with hs-crp, and the serum PAB level of the patients is negatively correlated with hs-crp, with a correlation coefficient R=-0.474. After t test of PAB, t=-3.32, p=0.003, it shows a statistical difference (figure 2A).

The level of PAB in the serum of patients is linearly correlated with CD4^+^T cells, and the level of PAB in the serum of patients is positively correlated with CD4^+^T cells, with a correlation coefficient R=0.467. After T test of PAB, T =3.08, p=0.004, it shows a statistical difference (figure 2C).

The level of PAB in the serum of the patients is linearly correlated with CD8^+^T cells, and the level of PAB in the serum of the patients is positively correlated with CD8^+^T cells, with a correlation coefficient R=0.613. After T test of PAB, T =4.526, p=0.000, it shows a statistical difference (figure 2D).

## 1. Discussion

This paper is the first reported clinical study in a non-Hubei study in China. There are several differences, compared with the clinical analysis results reported by Wuhan Jinyintan Hospital and Tongji Hospital Affiliated to Tongji Medical College of Huazhong University of Science and Technology (refer to as Tongji Hospital Affiliated hereafter). Only 2 of the 36 cases included in this study were assessed as critical on admission, while the rest of the cases were mainly of normal type and not severe.^[2][3]^ On admission, 50% of the patients had fever and cough as their first symptoms, mainly moderate and low fever, with few gastrointestinal symptoms. In this study, the lymphocyte absolute value reduction is only in 5 cases (13.9%), hypersensitive c-reactive protein increased in 19 (52.78%), significantly lower than the 69% and 93% reported by Tongji Hospital Affiliated s, or less than 35% and 86% of the Jinyintan Hospital reports respectively. That may be involved with the fact that this study is mainly based on the common type and mild type. The changes of lymphocyte absolute value and hypersensitive c-reactive protein in patients with the common and mild cases of the new coronavirus pneumonia are not as evident as those in the severe and critical cases, which can be easily ignored during clinical observation. In this study, we found a decrease in prealbumin in 15 cases (41.67%), and no previous studies have reported prealbumin changes. Prealbumin is usually used in clinical evaluation of the nutritional status of the body, and the half-life of serum prealbumin is short, so its changes may occur before albumin. In this study, there were only 5 patients with decreased albumin, all of which were with a slight decrease, but 14 patients (41.67%)showed decreased prealbumin. The pulmonary CT findings of the patients were basically consistent with previous reports. Twenty-two (61.11%) of the patients showed abnormal findings, with ground glass density shadows.

Our study suggests that patients with novel coronavirus pneumonia experience a decrease in t-cell subsets on admission, and in combination with clinical typing, we speculate that immune function may be suppressed or impaired in the early stage of the disease. As a commonly used indicator to evaluate nutritional status in clinical practice, prealbumin is often utilized to evaluate tumor and postoperative recovery ^[4]^. Domestic scholar CAI Chuanqi et al. ^[5]^ found that hypoalbuminemia is an independent risk factor for the severity of disease in elderly pneumonia patients. This study found that the albumin in the disease group was basically normal, while the prealbumin was significantly lower than that in the normal control group. We analyzed the relationship between prealbumin and CD3^+^T cells, CD4^+^T cells and CD8^+^T cells respectively. The regression model showed that there is a linear and positive correlation between the level of prealbumin in the serum of patients and CD3^+^T cells, CD4^+^T cells and CD8^+^T cells, with the strongest correlation among CD8^+^T cells. Proalbumin is an acute reactive protein synthesized by liver cells with a half-life of 19 days ^[6]^. In the early stage of the disease, proalbumin changes, which may be caused by the consumption of proalbumin by the body after infection with the virus, leading to the reduction of proalbumin in peripheral blood. Recent studies have found that inflammation is an effective inhibitor of protein synthesis, and it may also inhibit the synthesis of visceral prealbumin during the occurrence and development of the disease, leading to a low level of prealbumin^[7]^. Due to fever and infection, the body is in a state of high decomposition and high energy demand. If at the same time with lack of energy and loss of appetite, the patient cannot ensure sufficient food intake through the mouth to meet the needs of the body. Meanwhile if combined with basic diseases of other viscera, moderate or severe malnutrition can easily appear in pneumonia patients. Clinical studies have found that malnutrition rates in patients with severe pneumonia are as high as 60%, with immune dysfunction. Enteral nutrition or other forms of nutritional support are usually provided only to patients with severe pneumonia. Good nutritional status not only reflects the body’s immune function to a certain extent, but also may play an irreplaceable role in fighting the emerging inflammatory storm. Prealbumin can be used as an early warning indicator of immune function in patients with new coronavirus pneumonia. Through the analysis of the relationship between prealbumin and T cell subsets, the decreased value of prealbumin indicates that the patient’s cellular immune function is impaired, which plays a certain role in clinical condition monitoring.

According to the current outbreak reports, most of the patients are mild and normal type. In this study, we concluded that lymphocyte changes are not significant in the early stage of the disease in both normal and mild patients, and that changes in t-cell subsets and prealbumin might be beneficial for evaluating the disease.

## Data Availability

The datasets used and/or analyzed during the current study are available from the corresponding author on reasonable request.

## Notes

### Competing Interest Statement

The authors have declared no competing interest.

### Funding Statement

Funding information is not applicable.

